# Pausing methotrexate improves immunogenicity of COVID-19 vaccination in elderly patients with rheumatic diseases

**DOI:** 10.1101/2021.11.17.21266441

**Authors:** AN Arumahandi de Silva, LM Frommert, FN Albach, J Klotsche, V Scholz, LM Jeworowski, T Schwarz, A ten Hagen, J Zernicke, VM Corman, C Drosten, GR Burmester, R Biesen

## Abstract

**Objective:** To study the effect of methotrexate (MTX) and its discontinuation on the humoral immune response after COVID-19 vaccination in patients with autoimmune rheumatic diseases (AIRD).

**Methods:** In this retrospective study, neutralising SARS-CoV-2 antibodies were measured after second vaccination in 64 rheumatic patients on methotrexate therapy, 31 of whom temporarily paused medication without a fixed regimen. The control group consisted of 21 AIRD patients without immunosuppressive medication.

**Results:** MTX patients showed a significantly lower mean antibody response compared to AIRD patients without immunosuppressive therapy (71.8 % vs 92.4 %, p<0.001). For patients taking MTX, age correlated negatively with immune response (r=-0.49; p<0.001). All nine patients with antibody levels below the cut-off were older than 60 years. Patients who held MTX during at least one vaccination showed significantly higher mean neutralising antibody levels after second vaccination, compared to patients who continued MTX therapy during both vaccinations (83.1 % vs 61.2 %, p=0.001). This effect was particularly pronounced in patients older than 60 years (80.8 % vs 51.9 %, p=0.001). The impact of the time period after vaccination was greater than of the time before vaccination with the critical cut-off being 10 days.

**Conclusion:** MTX reduces the immunogenicity of SARS-CoV-2 vaccination in an age-dependent manner. Our data further suggest that holding MTX for at least 10 days after vaccination significantly improves the antibody response in patients over 60 years of age.

## INTRODUCTION

Until November 2021, SARS-CoV-2 had infected at least 250 million people worldwide and caused about 5 million deaths in a 23-month period ^1^. At the same time, enormous knowledge about SARS-CoV-2 and the related disease COVID-19 have been generated and the possibilities for prevention, diagnostics and treatments have improved remarkably.

Methotrexate (MTX) has been used for decades to treat a wide variety of immune-mediated diseases in oncology, rheumatology, dermatology, gastroenterology, and neurology. Following prednisolone, MTX is the most prescribed anti-inflammatory drug worldwide with 1 million MTX patients in the US alone ^2^.

Various immunosuppressants reduce the immune response after COVID-19 vaccination ^3^.. Although several research groups have recently described a reduced vaccination response under MTX ^4 5^, in some cohorts MTX had no negative influence ^6 7^. Most of these studies did not collect data on whether or not patients had paused MTX during vaccinations, although more than one-third of patients had modified their medication on their own or on the advice of their rheumatologist, according to a recent survey ^8^. The discontinuation of immunosuppressive medication can improve the vaccination response as recently shown for mycophenolate ^9^.

A reduced vaccination response under MTX was first described in 2016 for influenza vaccination ^10^. Follow-up data showed the increase in humoral immune response when pausing MTX two weeks before and after vaccination or only two weeks after vaccination ^11 12^. The time after and not before vaccination was decisive ^13^. However, data regarding MTX-hold during COVID-19 vaccination are still lacking, which is why current guidelines are based on experience with influenza vaccines, not considering mRNA based technology used for COVID-19 vaccinations. Although current guidelines by the American College of Rheumatology as well as the German Society for Rheumatology recommend holding MTX one to two weeks after COVID-19 vaccination ^14 15^, the EULAR does not recommend pausing MTX ^16^.

Therefore, our main objective was to study the effect of MTX and its discontinuation on the humoral immune response after COVID-19 vaccination in patients with autoimmune rheumatic diseases (AIRD). Secondary objective was to determine additional influencing factors on antibody response in these patients.

## METHODS

### Study design and participants

This is a retrospective sub-analysis of the VACCIMMUN Study, which is an observational cohort study among patients with autoimmune rheumatic diseases (AIRD) at the Charité Department for Rheumatology and Clinical Immunology in Berlin, Germany. Participants were recruited between April and September 2021 and had to meet the following inclusion criteria: age 18 years or older, AIRD diagnosis and vaccination with a COVID-19 vaccine authorised for use in Germany. For this analysis, only AIRD patients under MTX therapy were considered, receiving either only MTX or MTX combined with low dose prednisolone (defined as ≤ 5 mg/d), TNFα-Inhibitors, hydroxychloroquine, leflunomide, IL-17 or IL-12/23 inhibitors, since these immunosuppressive comedications are not known to have a remarkable impact on the immune response after vaccination ^15^. Additionally, AIRD patients who were vaccinated under no immunosuppressive therapy served as controls. Information regarding medical history including COVID-19 vaccination status and immunosuppressive therapy were provided directly by patients and additionally validated with medical records. At the time of blood drawing, patients were asked about their MTX intake schedule around vaccinations. The decision on continuing or holding MTX was made by the patient or the attending physician and was only observed in the study. Patients who reported to have changed their MTX-intake schedule resulting in an MTX-interval longer than 7 days around first or second vaccination were compared to patients who continued MTX therapy throughout both vaccinations. This study was ethically approved by the Regional Office for Health and Social Affairs Berlin, Germany (21/0098-IV E 13). All patients provided written informed consent.

### Laboratory analyses

Antibody response was measured predominantly about two weeks after the second dose of vaccination with maximum range from 11 to 112 days. Neutralising antibody levels were assessed using a surrogate virus neutralisation test (cPass Neutralisation, Medac GmbH, Wedel, Germany)^17^. Following the manufacturer’s protocol, patients who reached inhibition rates greater than or equal to 30 % were considered to have demonstrated a SARS-CoV-2 specific humoral response and are further defined as responders, while patients with inhibition rates <30 % are defined as non-responders. Additionally, IgG antibodies against nucleocapsid, receptor binding domain (RBD), full spike and the S1 domain of the spike protein were tested using SeraSpot® Anti-SARS-CoV-2 IgG microarray-based immunoassay (Seramun Diagnostica GmbH, Heidesee, Germany) and served here for further validation purposes. Hence, all calculations were additionally performed using anti-RBD-IgG levels and can be found in the supplements. The threshold for reactivity for anti-SARS-CoV-2 IgG levels was set at >1.00 Signal/Cut-off (S/CO) in accordance with manufacturer’s protocol.

### Statistical analysis

Descriptive statistics included mean with standard deviation and absolute and relative frequencies. The exact unconditional z-pooled test^18^ and Chi^2^ test were applied for binary and categorical data and the unpaired t test with Welch’s correction for continuously distributed variables to perform hypotheses tests for group differences, as appropriate. The likelihood of response to vaccination was modelled by a Poisson generalized linear model with robust error variances and log link function including the covariates age, gender, MTX monotherapy, MTX in combination with prednisolone, MTX in combination with other DMARDs ± prednisolone, MTX-hold and vaccine interval as suggested by Zou ^19^. These covariates were selected based on the theoretical assumption that they could affect vaccination success and on the results of the univariate analysis. The association between antibody results (dependent variables anti-RBD-IgG concentrations or neutralising capacity) and the covariates age, gender, MTX monotherapy, MTX in combination with prednisolone and MTX in combination with other DMARDs ± prednisolone, MTX-hold, vaccine interval and timing and duration of MTX-hold was estimated by a linear regression model. The unstandardized and standardized beta coefficients were calculated for linear regression analyses in order to compare the strengths of association between parameters. The area under curve was calculated after fitting a logistic regression model to provide a measure of strengths of association for dichotomous outcomes. The Youden index was used to estimate thresholds for age and time of MTX break before and after vaccination from receiver-operating characteristics. Statistical analyses were performed using Graphpad Prism 9.2.0, R 4.1.2 and STATA 12.1.

### Patient and public involvement

This study aimed to provide evidence for future recommendations due to questions asked regarding MTX intake by patients and physicians. However, patients and the public were not directly involved in process of designing.

## RESULTS

### Patient characteristics

Of 73 eligible patients receiving MTX, nine were excluded due to unacceptable immunosuppressive comedication, irregular medication regimens and unclassifiable MTX-hold. The final cohort consisted of 64 AIRD patients taking MTX (mean age 61 years, 70.3 % women) and 21 rheumatic patients who did not receive any kind of immunosuppressive therapy as a control group (mean age 61, 90.5 % women). Detailed clinical characterisation is given in supplementary table 1. Patients in the no-therapy group were of similar age and BMI, but more often female. They were less often diagnosed with rheumatoid arthritis and more often with systemic sclerosis.

Of 64 MTX patients, 31 patients reported to have held MTX for at least one vaccination (MTX-hold) while 33 patients had continued their MTX therapy without any interruption (MTX continued, table 1). Blood sampling occurred slightly earlier in the MTX-hold group than in the MTX continued group. There were no significant differences between these two groups regarding age, BMI, distribution of sex, vaccination regimes, diagnoses and immunosuppressive comedications (table 1).

**Table 1:**
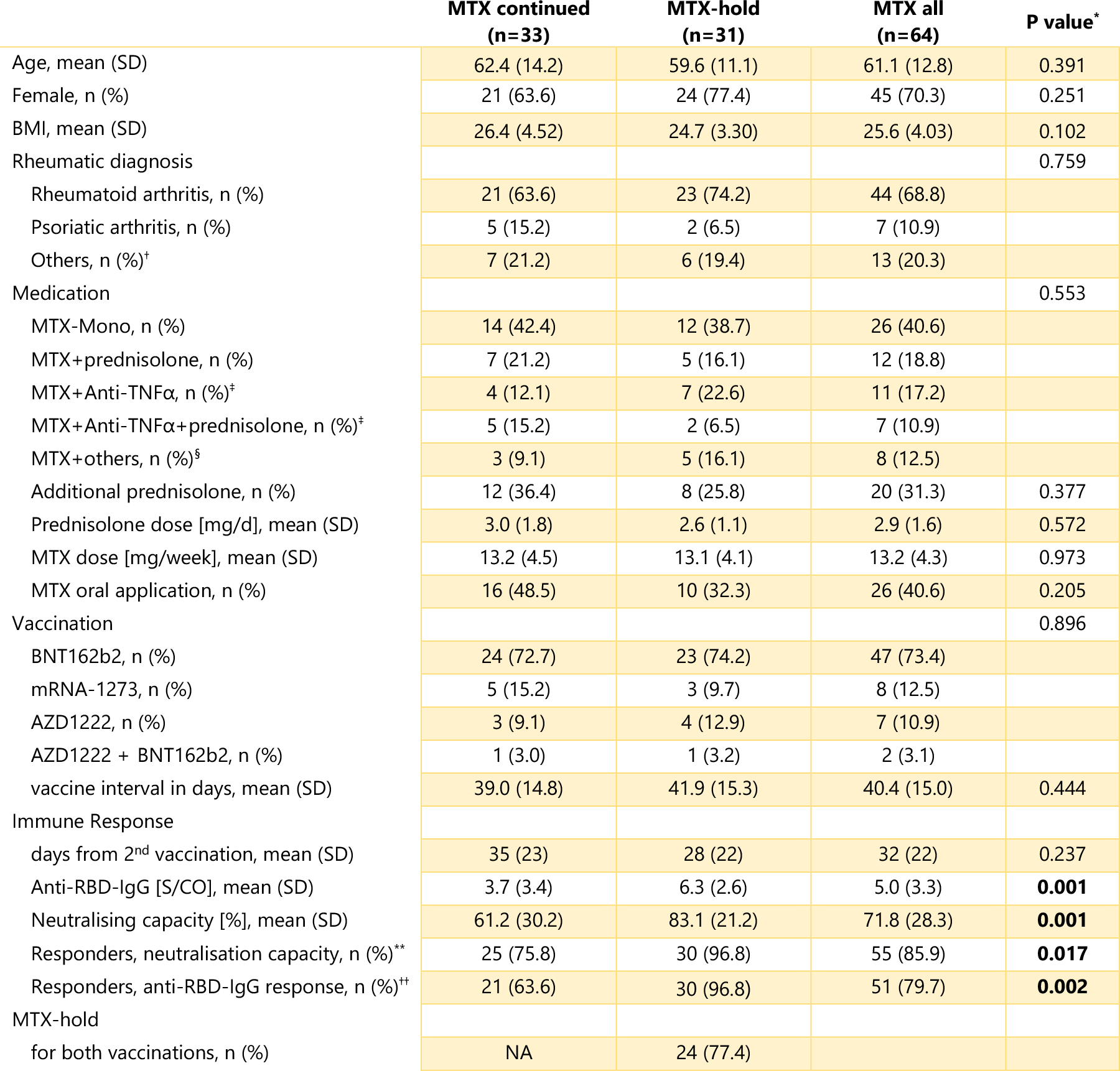
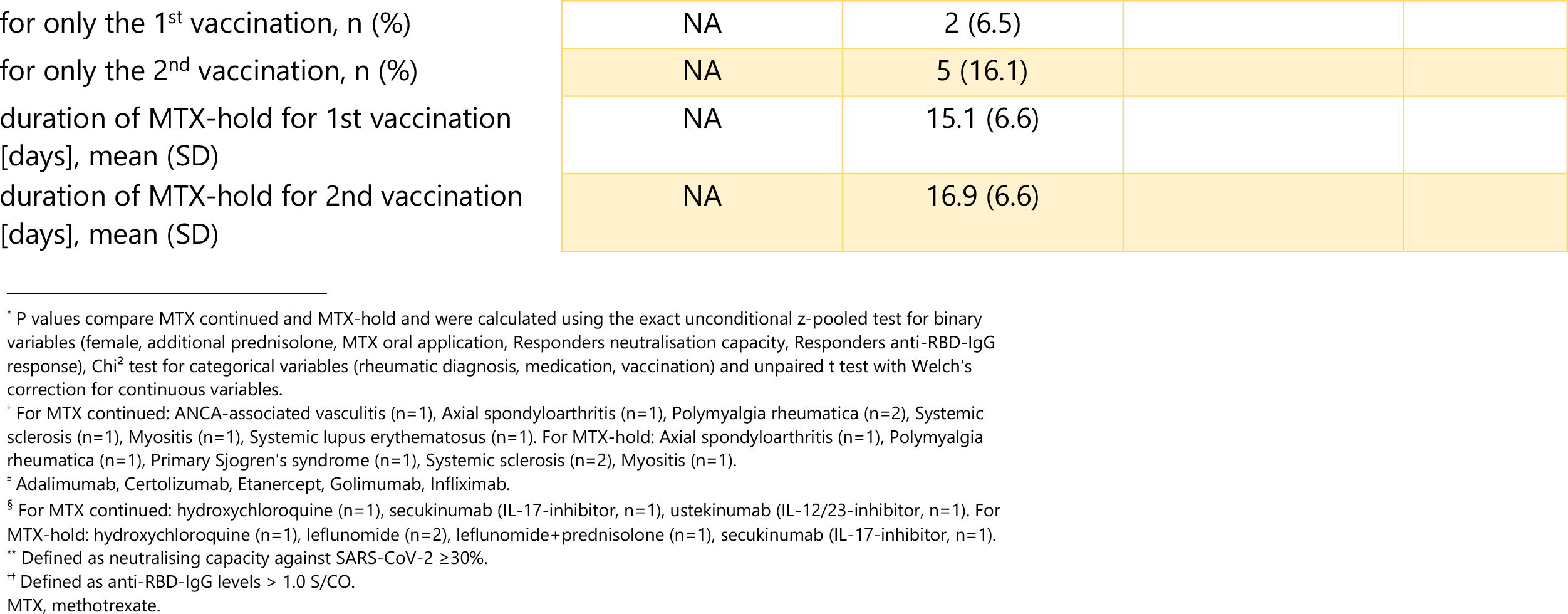
Characteristics of MTX patients who held and continued MTX

### MTX reduces vaccination response

AIRD patients without immunosuppressive therapy showed a significantly higher neutralising capacity (mean 92.4 %, SD: 8.6) than AIRD patients taking MTX (mean 71.8 %, SD: 28.3, p< 0.001, figure 1, supplementary figure 1 for anti-RBD-IgG). This was still the case after adjusting for the possible confounders gender, age, vaccine regime and vaccine interval, AIRD diagnosis and duration from second vaccination to blood draw in a logistic regression (for neutralising capacity: beta = -19.5, 95 % CI: -31.4 ; -7.7, p=0.002; for anti-RBD-IgG: beta = -1.61, 95 % CI: -3.03 ; -0.18, p=0.028). None of the patients without immunosuppressive therapy were classified as non-responders (defined by neutralisation activity < 30 %), compared to 14.1 % (n=9) among MTX patients. Taking patients without immunosuppressive therapy in our cohort as a reference group for a typical antibody response after vaccination, the threshold for a not-altered inhibition rate could be set at 87.6 % (AUC 0.75, Youden index 49.9). Accordingly, 38 of 64 MTX patients (59.4 %) demonstrated a lower antibody response after two vaccinations compared to an untreated group of AIRD patients.

**Figure 1:**
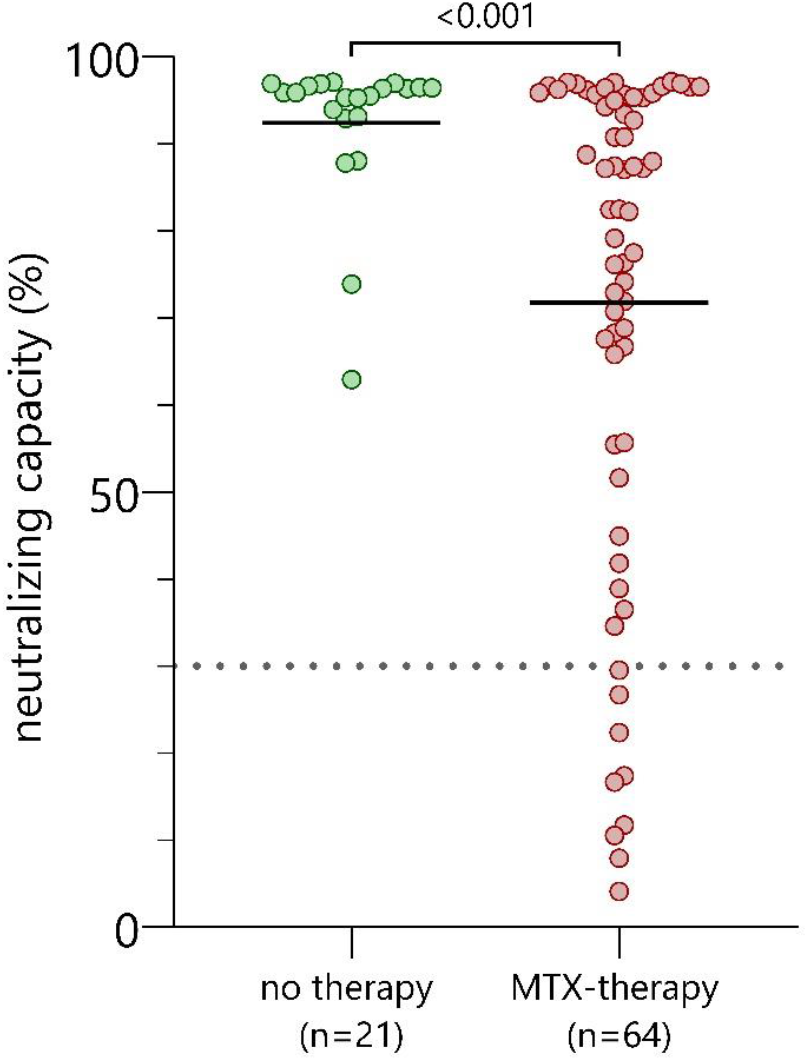
Comparison of neutralising capacity in AIRD patients without immunosuppression and with MTX therapy. Neutralising capacity measured using surrogate virus neutralisation test after second vaccination in MTX-patients (n=64) represented by red dots vs. AIRD patients who were under no immunosuppressive therapy during both vaccinations (n=21) represented by green dots. P values were calculated using the parametric unpaired t test with Welch’s correction.

### Factors influencing antibody response in MTX patients

To identify factors influencing the antibody response under MTX we compared COVID-19 vaccination responders (n=55, 85.9 %) and non-responders (n=9, 14.1 %) defined by neutralisation activity. Both groups were comparable in BMI, vaccine type, MTX application form, additional prednisolone intake, time of blood draw and immunosuppressive comedication (table 2). Dosage of MTX was not significantly associated with vaccination success (Spearman rank correlation, r=-0.02, p value=0.867). However, a higher neutralisation capacity was significantly associated with young age, MTX-hold and female gender in univariate analysis (table 2) and multivariable analysis (table 3). If classification into responders and non-responders was based on anti-RBD-IgG results, 13 patients fall into the non-responder group. While the effect of age and MTX-hold were still significant using anti-RBD-IgG levels, this was not the case for gender (supplementary table 2, table 3). A longer vaccine interval was associated with an adequate humoral response to vaccination in our cohort (significant in t test for neutralising capacity and anti-RBD-IgG levels; only significant in multivariable analysis for anti-RBD-IgG levels). In the following, we will analyse the effect of age and MTX-hold in more detail.

**Table 2:**
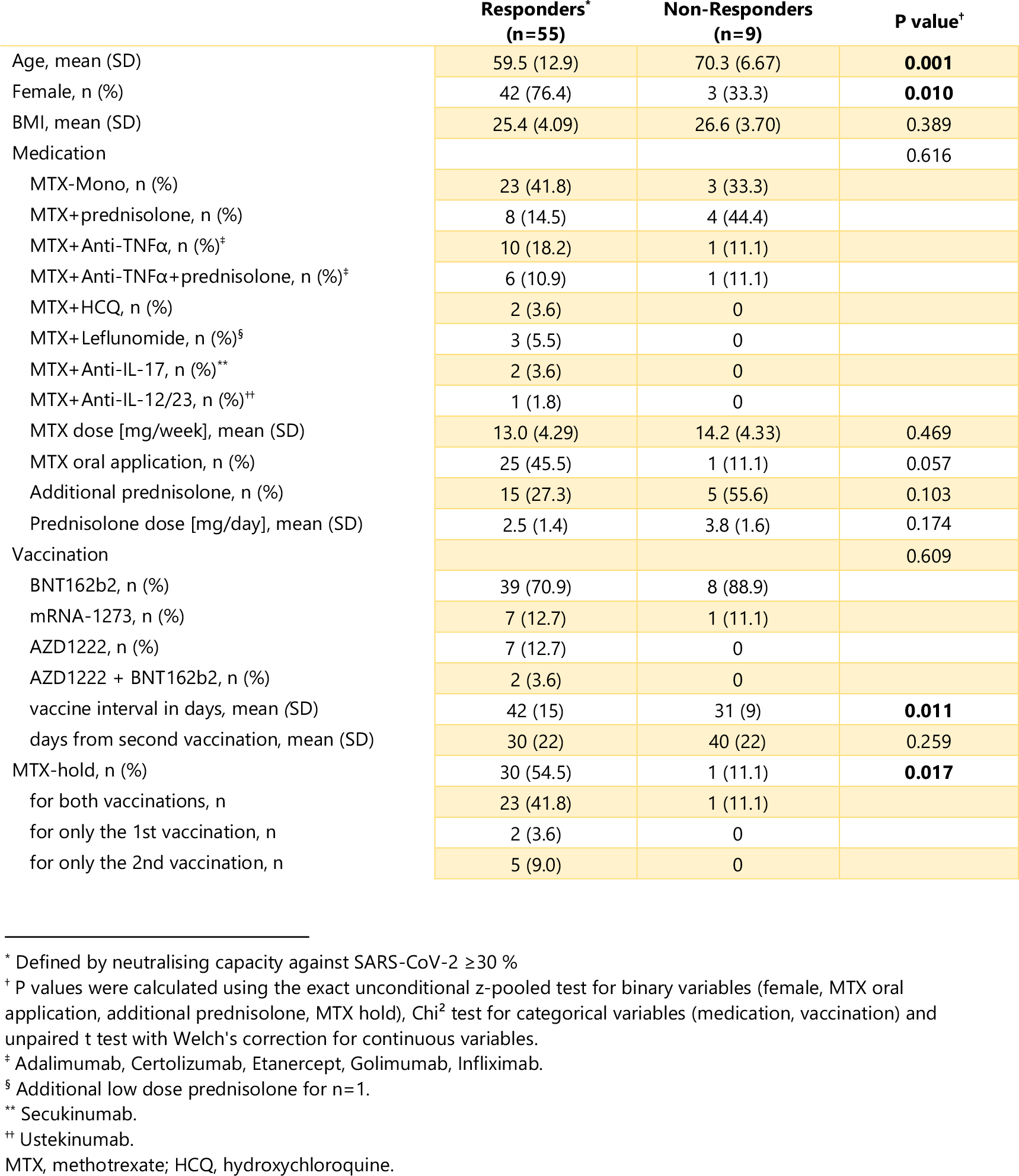
Comparison of vaccination responders and non-responders among AIRD-patients taking MTX

**Table 3:** Association of neutralising capacity and Anti-RBD-IgG concentration with selected covariates in univariate and multivariable analyses (n=64).

|  | univariate analysis |  |  |  | multivariable analysis |  |  |
| --- | --- | --- | --- | --- | --- | --- | --- |
|  | RR †† | p value | 95%CI | AUC | RR †† | p value | 95%CI |
| **Outcome: Anti-RBD-IgG concentration >1 S/CO** |  |  |  |  |  |  |  |
| Female | 1.18 | 0.280 | 0.88 ; 1.58 | 0.60 | 1.23 | 0.125 | 0.94 ; 1.62 |
| Age in years†† | **0.93** | **<0.001** | **0.89 ; 0.97** | **0.89** | **0.94** | **0.001** | **0.90 ; 0.97** |
| MTX monotherapy | 1.00 |  |  | 0.63 | 1.00 |  |  |
| MTX + prednisolone | 0.79 | 0.284 | 0.51 ; 1.22 |  | 0.86 | 0.415 | 0.60 ; 1.24 |
| MTX combination ± prednisolone | 1.05 | 0.687 | 0.84 ; 1.30 |  | 1.04 | 0.693 | 0.86 ; 1.25 |
| MTX dose in mg | 0.99 | 0.688 | 0.97 ; 1.02 | 0.54 |  |  |  |
| MTX hold | **1.39** | **0.006** | **1.10 ; 1.76** | **0.74** | **1.27** | **0.020** | **1.04 ; 1.56** |
| Vaccine interval | **1.006** | **0.016** | **1.001 ; 1.010** | **0.75** | **1.004** | **0.024** | **1.0006 ; 1.008** |
| **Outcome: neutralisation capacity ≥30%** |  |  |  |  |  |  |  |
| Female | 1.36 | 0.055 | 0.99 ; 1.87 | 0.72 | **1.43** | **0.012** | **1.08 ; 1.90** |
| Age in years†† | **0.96** | **0.008** | **0.92 ; 0.99** | **0.77** | **0.96** | **0.018** | **0.93 ; 0.99** |
| MTX monotherapy | 1.00 |  |  | 0.68 | 1.00 |  |  |
| MTX + prednisolone | 0.75 | 0.194 | 0.49 ; 1.15 |  | 0.75 | 0.099 | 0.53 ; 1.06 |
| MTX combination ± prednisolone | 1.04 | 0.641 | 0.87 ; 1.25 |  | 0.98 | 0.838 | 0.84 ; 1.15 |
| MTX dose in mg | 0.99 | 0.452 | 0.97 ; 1.01 | 0.58 |  |  |  |
| MTX hold | **1.28** | **0.019** | **1.04 ; 1.57** | **0.72** | **1.17** | **0.039** | **1.00 ; 1.38** |
| Vaccine interval | 1.002 | 0.235 | 0.999 ; 1.006 | 0.63 | 1.001 | 0.423 | 0.998 ; 1.004 |

AUC = Area under Curve; 95%CI = 95 % confidence interval; RR = relative risk.
†† Relative risk was estimated by a Poisson generalized linear model with robust error variances and log link function in univariate and multivariable analyses according to Zou <sup>19</sup>.
† relative risk for increase by 5 years

### Effect of MTX-hold and age

Patients who had changed their MTX intake schedule for at least one vaccination showed a significantly higher antibody response than patients who continued their MTX intake (p=0.001, figure 2A, supplementary figure 2A for anti-RBD-IgG). Mean neutralisation was 61.2 % for patients who continued their therapy and 83.1 % for patients who held MTX (table 1). There was only one non-responder (3.2 %) in the MTX-hold group, while there were eight non-responders (24.2 %) in the MTX continued group. The effect of pause persisted in patients with MTX monotherapy, indicating that this effect cannot be explained by the existing comedication (supplementary figure 3).

**Figure 2:**
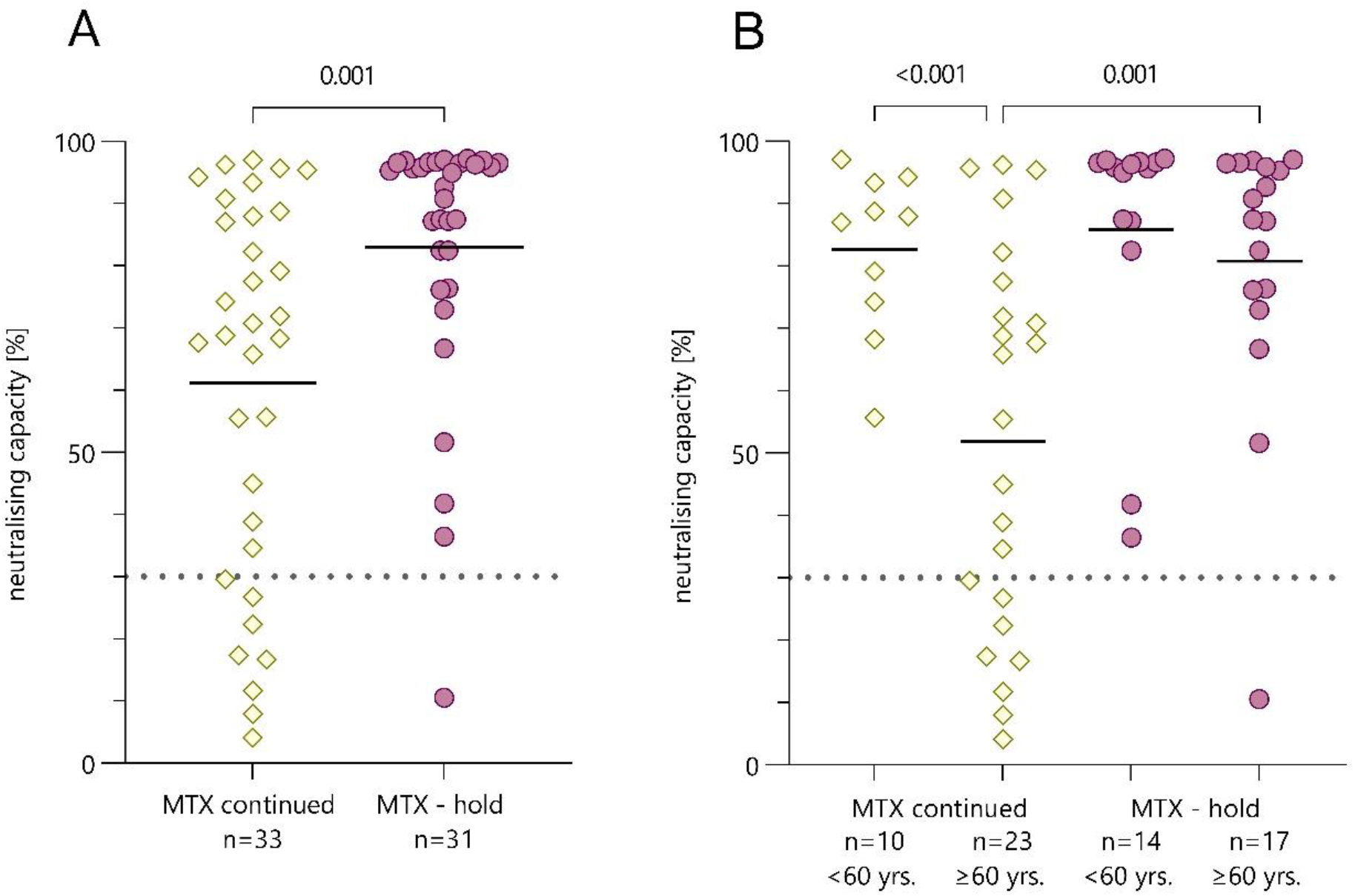
Comparison of AIRD patients who continued or held their MTX during the COVID-19 vaccination. (A) Neutralising capacity measured using surrogate virus neutralisation test compared between patients who held MTX during vaccination (n=33) and patients who continued MTX-therapy (n=31). (B) Neutralising capacity differentiated by age groups <60 years and ≥60 years. P values were calculated using the parametric unpaired t test with Welch’s correction. Dotted line marks the cut-off value following manufacturer’s protocol (≥30 %). Yellow squares represent patients who continued MTX-therapy, purple dots represent patients who held MTX for at least one vaccination.

Vaccination response correlated significantly with age (Spearman rank correlation, -0.49, p<0.001, figure 3, supplementary figure 4 for anti-RBD-IgG). No patient younger than 60 years was classified a non-responder which is why we further distinguished the MTX hold and continued groups into patients older and younger than 60 years of age (figure 2B, supplementary figure 2B for anti-RBD-IgG). Considering only patients who continued their MTX intake, patients ≥ 60 years of age (mean 51.9 %) had a 30.7 percentage points lower mean inhibition rate than patients < 60 years (mean 82.6 %). Vice versa, neutralisation levels were 28.9 percentage points higher in patients older than 60 years who held MTX (mean 80.8 %) compared to those who continued MTX (mean 51.9 %). In contrast, when regarding patients under 60 years there were no significant differences in neutralisation rates between patients who held or continued MTX therapy.

**Figure 3:**
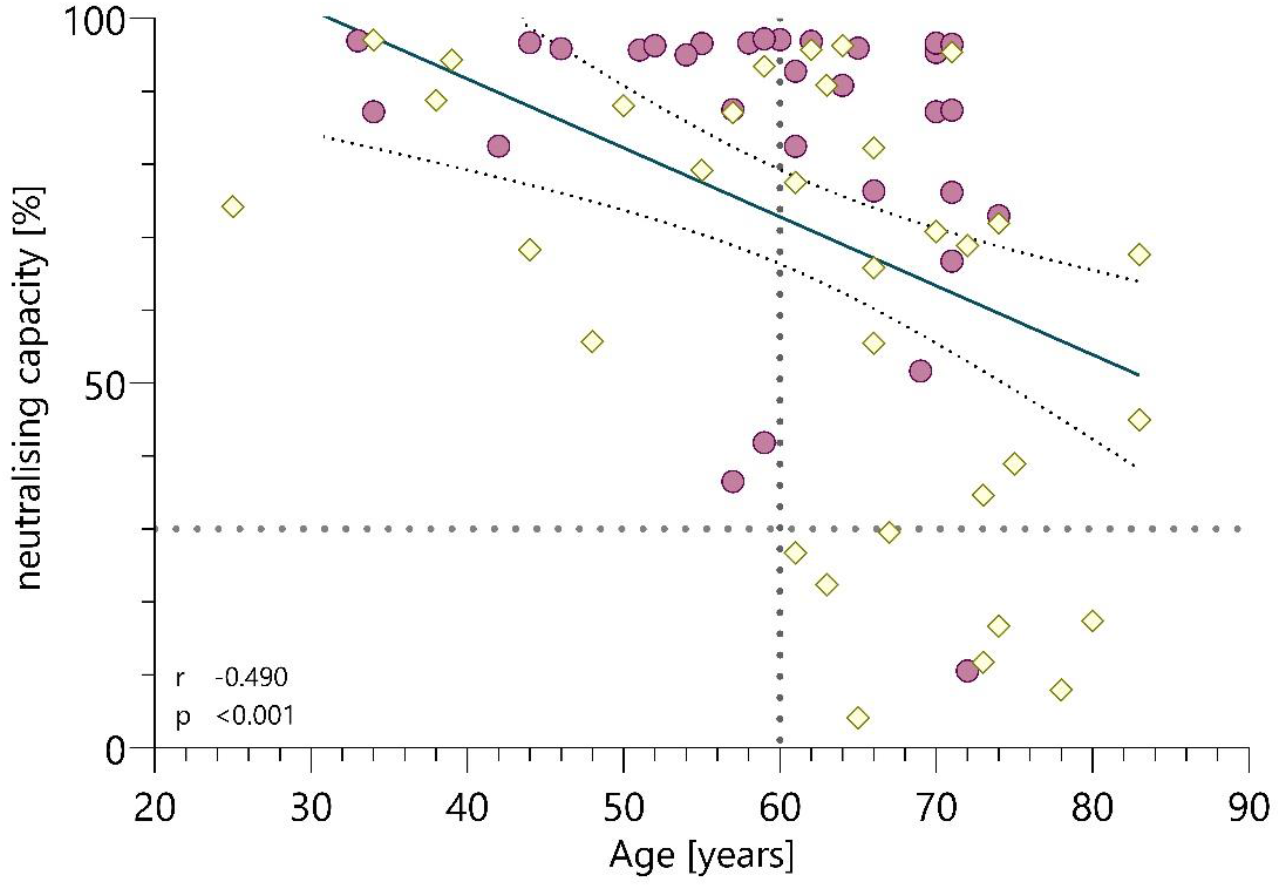
Correlation of Age and neutralising capacity measured using surrogate virus neutralisation test. Purple dots represent patients who held MTX during vaccination (n=31), yellow squares represent patients who continued MTX-therapy (n=33). Neutralising antibodies were measured using a surrogate virus neutralisation test. Dotted lines mark the cut-off value following manufacturer’s protocol (≥30 %) and the cut-off age used for further analysis at 60 years. P value and correlation coefficient were calculated using the Spearman Rank correlation.

### Effect of timing and duration of MTX-hold

In the following we considered all 64 patients and analysed the MTX-interval at the time of vaccination, which was defined by the time between last MTX intake and vaccination (time before vaccination = T_BV_) and the time between vaccination and re-intake of MTX (time after vaccination = T_AV_, figure 4). One patient could not recall on which day MTX was taken and was therefore not considered for calculations of T_BV_ and T_AV_. We found that the duration of the MTX-interval (T_BV_ + T_AV_) significantly correlates with neutralising capacity (Spearman rank correlation, r = 0.47, p< 0.001). We further analysed which of these time periods is most likely to determine antibody response. By using linear regression analysis, we found time after vaccination (T_AV_) to be highly significant for adequate neutralisation rate and anti-RBD-IgG concentration in the elderly, but not for younger patients (table 4). Here, ten days between vaccination and MTX re-intake (T_AV_) were determined as the critical cut-off based on the Youden index from receiver operating characteristic curve.

**Figure 4:**
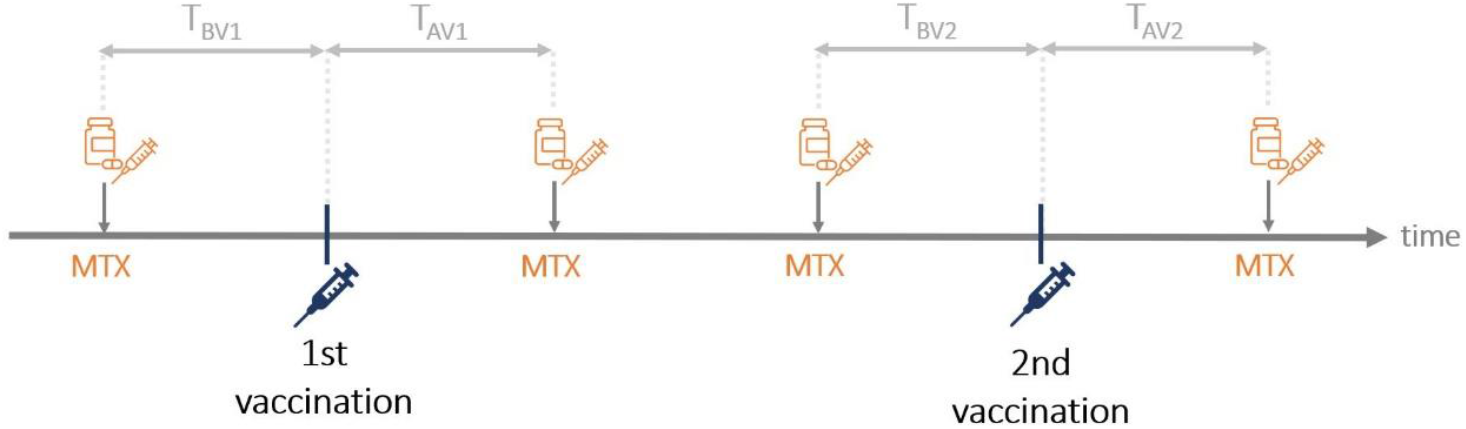
Visualization of analysed time intervals. Time between MTX-intakes and COVID-19 vaccinations were assessed for each vaccination and added together to receive the total time before vaccinations (TBV=TBV1+TBV2) and after vaccinations (TAV=TAV1+TAV2). The MTX-interval was defined as the total durations between two MTX intakes at the time of vaccination (TAV+TBV).

**Table 4:** Association of Neutralising capacity and Anti-RBD-IgG concentration with MTX-intake timing using linear regression analysis (n=64)^*^

|  | All patients<sup>†</sup> |  |  |  | Patients < 60 years<sup>†</sup> |  |  |  | Patients ≥ 60 years<sup>†</sup> |  |  |  |
| --- | --- | --- | --- | --- | --- | --- | --- | --- | --- | --- | --- | --- |
|  | β | P value | 95% CI | β<sub>st</sub> | β | P value | 95% CI | β<sub>st</sub> | β | P value | 95% CI | β<sub>st</sub> |
| **neutralisation capacity** |  |  |  |  |  |  |  |  |  |  |  |  |
| T<sub>BV</sub> | 0.00 | 0.976 | -0.20 ; 0.19 | 0.00 | -0.05 | 0.749 | -0.37 ; 0.27 | -0.06 | 0.04 | 0.807 | -0.26 ; 0.33 | 0.03 |
| T<sub>AV</sub> | **0.19** | **0.005** | **0.06 ; 0.32** | **0.30** | 0.09 | 0.301 | -0.09 ; 0.28 | 0.20 | **0.24** | **0.012** | **0.06 ; 0.43** | **0.32** |
| T<sub>BV</sub> ≥ 10 days | -0.85 | 0.367 | -2.73 ; 1.02 | -0.08 | -0.36 | 0.789 | -3.12 ; 2.40 | -0.05 | -1.98 | 0.094 | -4.31 ; 0.35 | -0.13 |
| T<sub>AV</sub> ≥ 10 days | **2.00** | **0.008** | **0.53 ; 3.47** | **0.27** | -0.22 | 0.833 | -2.34 ; 1.91 | -0.04 | **2.91** | **0.001** | **1.29 ; 4.53** | **0.36** |
| T<sub>BV</sub> + T<sub>AV</sub> | **0.13** | **0.035** | **0.01 ; 0.25** | **0.25** | 0.04 | 0.626 | -0.13 ; 0.22 | 0.11 | **0.18** | **0.039** | **0.01 ; 0.35** | **0.29** |
| **anti-RBD-IgG** |  |  |  |  |  |  |  |  |  |  |  |  |
| T<sub>BV</sub> | 0.80 | 0.244 | -0.56 ; 2.15 | 0.09 | 0.47 | 0.575 | -1.24 ; 2.17 | 0.10 | 1.26 | 0.236 | -0.86 ; 3.39 | 0.12 |
| T<sub>AV</sub> | **1.02** | **0.016** | **0.19 ; 1.84** | **0.19** | 0.11 | 0.739 | -0.56 ; 0.78 | 0.04 | **1.63** | **0.036** | **0.12 ; 3.14** | **0.22** |
| T<sub>BV</sub> ≥ 10 days | -1.86 | 0.777 | -14.96 ; 11.24 | -0.02 | 0.49 | 0.950 | -15.49 ; 16.47 | 0.01 | -6.43 | 0.323 | -19.43 ; 6.57 | -0.05 |
| T<sub>AV</sub> ≥ 10 days | **13.70** | **0.004** | **4.60 ; 22.79** | **0.21** | -2.53 | 0.583 | -11.96 ; 6.90 | -0.07 | **20.03** | **0.005** | **6.57 ; 33.50** | **0.26** |
| T<sub>BV</sub> + T<sub>AV</sub> | **0.95** | **0.003** | **0.33 ; 1.58** | **0.22** | 0.21 | 0.487 | -0.41 ; 0.84 | 0.09 | **1.51** | **0.019** | **0.26 ; 2.77** | **0.26** |

\* One patient who did not hold MTX could not recall on which exact day MTX was taken and was therefore only considered for calculations of T<sub>BV</sub> + T<sub>AV</sub> (=7 days).
<sup>†</sup> adjusted for female, age, MTX monotherapy, MTX + prednisolone, MTX combination ± prednisolone, vaccine interval.
<sup>‡</sup> adjusted for female, MTX monotherapy, MTX + prednisolone, MTX combination ± prednisolone, vaccine interval.
β (unstandardized beta coefficient); β<sub>st</sub> (standardized beta coefficient); TBV (time before vaccination), time between last MTX-intake and vaccination; TAV (time after vaccination), time between vaccination and re-intake of MTX; TBV+TAV, MTX-interval at the time of vaccination

## DISCUSSION

Our study found a reduced COVID-19 vaccination response in MTX patients, demonstrates the effect of age and provides first data on the effect of MTX-hold around COVID-19 vaccinations.

Using neutralising capacity and the manufacturer’s cut-off, we found a slightly higher rate of vaccination responders among patients taking MTX (85.9 %) than previously reported (47 – 72 %) ^4 5^. Using ROC analysis and an untreated control group, we determined an adapted cut-off value and found adequate immune response in only 40.6 % of MTX patients. Hence, we confirmed the observations from previous studies that the antibody response is reduced under MTX therapy ^4 5^. In contrast, others described no effect of MTX on vaccination response ^6 7^. These varying results may be due to a lower effect size of MTX on vaccination response compared to other immunosuppressive therapies such as rituximab or mycophenolate, different test systems and statistical analyses used and other influencing factors such as age and pausing of MTX therapy.

We determined young age, MTX-hold and longer vaccine interval as the main factors improving antibody response after vaccination. The negative influence of age on vaccination response was already known ^20 21^. However, the consideration of age was not yet differentiated in previous studies investigating immune response under MTX therapy. Therefore our data allows the assumption that continuous MTX intake and old age are potentiating negative factors. The positive effect of a longer vaccine interval on humoral immune response is in line with previously published works ^22 23^. These results were statistically significant in t test for both antibody testing systems, but in the generalized linear model only for anti-RBD-IgG levels. This discrepancy is likely due to the higher statistical power of the t test.

Patients who held MTX for at least one vaccination had a significantly higher immune response than those who continued MTX, which has not yet been described for COVID-19 vaccination. Nevertheless, our findings are in line with studies by Park et al. investigating the effect of MTX-hold on the immune response to influenza vaccination ^11^. More detailed analysis showed that time after vaccination is crucial, which was also described by Park et al. who recommended a MTX discontinuation of two weeks after influenza vaccination ^12 13^. In our study, we found a minimum time of 10 days after vaccination to be critical for immune response in patients ≥ 60 years. Additionally, the positive effect of MTX-hold was only statistically significant for patients 60 years or older. An effect also in younger patients might be observed in a larger cohort.

A strength of our study was that we validated all our neutralisation test results with an additional test system measuring anti-RBD-IgG levels. The latter defined four more patients as non-responders compared to the neutralisation test. This small number of conflicting test results is to be expected when using different test systems. The uneven distribution of gender among patients who had conflicting test results caused our analyses to suggest a significant influence of gender on the neutralisation result. This may be due to a statistical artifact and the effect of gender should be interpreted with caution.

This study has limitations. Since data regarding the MTX intake schedule during vaccination was assessed retrospectively, recall bias cannot be excluded. Due to our small sample size, we had to limit factors in the multivariable logistic regression modelling, which may lead to bias and residual confounding. For instance, confounding due to duration from vaccination to blood sampling, disease activity or AIRD diagnosis cannot with certainty be excluded in our analyses. We did not assess disease activity and safety of pausing MTX in our cohort, but current data do not indicate a significantly higher flare occurrence or disease activity in association with MTX discontinuation of two weeks ^24^. Also, T-cell-response was not part of our study design. However, according to current studies, it can be assumed that measuring humoral vaccination response is an adequate mean to determine vaccine immunogenicity ^25^ and that higher antibody levels correlate with a better clinical outcome ^26 27^. To address these limitations, a randomised controlled clinical trial to generate evidence for optimal management of MTX in COVID-19 vaccinations should be performed.

In conclusion, we present real-world data that is of clinical relevance regarding ongoing booster vaccinations. We determined age and MTX-hold as the main factors influencing antibody response during SARS-CoV-2 vaccinations and both aspects should be considered when discussing MTX regimens. Our data suggest that, if possible, patients older than 60 years of age should hold MTX for at least 10 days after receiving a COVID-19-vaccination.

## Key messages

### What is already known about this subject?

- Patients receiving methotrexate (MTX) have a reduced immune response after COVID-19 vaccination and holding MTX has shown to increase the immunogenicity after influenza vaccination.
- Yet, no previous studies have analysed the effect of MTX-hold for COVID-19 vaccination.

### What does this study add?

- This study identified old age (≥ 60 years), short vaccine interval and MTX continuation as critical factors for an inadequate antibody response.
- We found a minimum of 10 days between vaccination and re-intake of MTX as the critical threshold to increase immunogenicity for patients ≥ 60 years of age.

### How might this impact on clinical practise or future developments?

- In regard of ongoing booster vaccinations, our data suggest that especially older MTX patients should hold MTX for at least 10 days after receiving a COVID-19 vaccination.

## Data Availability

Data are available on reasonable request.
All data relevant to the study are included in the article or uploaded as online supplemental information.

## Acknowledgements

We would like to thank Tanja Braun and Vera Höhne-Zimmer for their support in obtaining the ethics vote and for their organisational support.

## Contributors

All authors contributed to the acquisition, analysis or interpretation of data and critical revision of the manuscript for important intellectual content. RB had full access to all the data in the study and takes responsibility for the integrity of the data and the accuracy of the data analysis. FA, JZ, GB, RB were involved in the study design. Sample collection was done by AS, LF, FA, VS, AH, JZ. Experiments and data analysis were performed by AS, LF, FA, JK, LJ, TS, JZ, VC, CD, GB and RB. AS, LF, FA and RB were responsible for tables and figures. Data interpretation was done by all authors. Statistical analyses were done by AS, LF, FA, JK and RB. Writing of the manuscript were performed by AS, LF, FA, JK, GB and RB. All authors were involved in critical proof reading of the manuscript.

## Patient consent

All patients provided written informed consent.

## Ethics approval

The Berlin State Office for Health and Social Affairs (Turmstrasse 21, 10559 Berlin, Germany) has approved this study under file number 21/0098-IV E 13.

## Data sharing statement

Data are available on reasonable request. All data relevant to the study are included in the article or uploaded as online supplemental information.

## Tables

**Supplementary table 1:** Characteristics of MTX-patients and control-group

|  | MTX-therapy<br>(n=64) | No therapy<br>(n=21) | All<br>(n=85) | P value* |
| --- | --- | --- | --- | --- |
| Age, mean (SD) | 61 (13) | 61 (14) | 61 (13) | 0.899 |
| Female, n (%) | 45 (70.3) | 19 (90.5) | 64 (75.3) | 0.069 |
| BMI, mean (SD) | 25.6 (4.0) | 23.8 (3.9) | 25.1 (4.1) | 0.088 |
| Rheumatic diagnosis |  |  |  | **<0.001** |
| Rheumatoid arthritis, n (%) | 44 (68.6) | 3 (14.3) | 47 (55.3) |  |
| Psoriatic arthritis, n (%) | 7 (10.9) | 2 (9.5) | 9 (10.6) |  |
| Systemic sclerosis, n (%) | 3 (4.7) | 8 (38.1) | 11 (12.9) |  |
| Others, n (%)<sup>†</sup> | 10 (15.6) | 8 (38.1) | 18 (21.2) |  |
| Vaccination |  |  |  | 0.542 |
| BNT162b2, n (%) | 47 (73.4) | 16 (76.2) | 63 (74.1) |  |
| mRNA-1273, n (%) | 8 (12.5) | 2 (9.5) | 10 (11.8) |  |
| AZD1222, n (%) | 7 (10.9) | 1 (4.8) | 8 (9.4) |  |
| AZD1222 + mRNA, n (%)<sup>‡</sup> | 2 (3.1) | 2 (9.5) | 4 (4.7) |  |
| vaccine interval in days, mean (SD) | 40.4 (15.0) | 38.6 (21.4) | 40.0 (16.7) | 0.725 |
| Immune response |  |  |  |  |
| days from 2<sup>nd</sup> vaccination, mean (SD) | 32 (22) | 31 (20) | 32 (22) | 0.807 |
| Anti-RBD-IgG [S/CO], mean (SD) | 5.0 (3.3) | 6.8 (2.0) | 5.4 (3.1) | **0.004** |
| Neutralising capacity [%], mean (SD) | 71.8 (28.3) | 92.4 (8.6) | 76.9 (26.4) | **<0.001** |
| Responders, n (%)<sup>§</sup> | 55 (85.9) | 21 (100) | 76 (89.4) | 0.076 |

\* P values were calculated using the exact unconditional z-pooled test for binary variables (female, responders), Chi<sup>2</sup> test for categorical variables (rheumatic diagnosis, vaccination) and unpaired t test with Welch's correction for continuous variables.
<sup>†</sup> For MTX group: ANCA-associated vasculitis (n=1), Axial spondyloarthritis (n=2), Polymyalgia rheumatica (n=3), Primary Sjogren's syndrome (n=1), Myositis (n=2), Systemic lupus erythematosus (n=1).
For no therapy group: Axial spondyloarthritis (n=2), Polymyalgia rheumatica (n=1), Primary Sjogren's syndrome (n=2), Systemic lupus erythematosus (n=1), Familial Mediterranean fever (n=1), peripheral spondyloarthritis (n=1).
<sup>‡</sup> MTX group: AZD1222 + BNT162b2 (n=2). No-therapy-group: AZD1222 + BNT162b2 (n=1), AZD1222 + mRNA-1273 (n=1).
<sup>§</sup> Defined by neutralising capacity against SARS-CoV-2 $\geq 30$ %. MTX, methotrexate.

**Supplementary table 2:** Comparison of vaccination responders and non-responders (anti-RBD-IgG levels)

|  | **Responders\***<br>**(n=51)** | **Non-Responders**<br>**(n=13)** | **P value†** |
| --- | --- | --- | --- |
| Age, mean (SD) | 58 (12) | 73 (7) | **<0.001** |
| Female, n (%) | 38 (74.5) | 7 (53.8) | 0.163 |
| BMI, mean (SD) | 25.3 (3.9) | 26.5 (4.7) | 0.395 |
| Medication |  |  | 0.524 |
| MTX-Mono, n (%) | 22 (43.1) | 4 (30.8) |  |
| MTX+prednisolone, n (%) | 7 (13.7) | 5 (38.5) |  |
| MTX+Anti-TNFα, n (%)‡ | 9 (17.6) | 2 (15.4) | |
| MTX+Anti-TNFα+prednisolone, n (%)‡ | 5 (9.8) | 2 (15.4) | |
| MTX+HCQ, n (%) | 2 (3.9) | 0 |  |
| MTX+Leflunomide, n (%)§ | 3 (5.9) | 0 |  |
| MTX+Anti-IL-17, n (%)** | 2 (3.9) | 0 |  |
| MTX+Anti-IL-12/23, n (%)†† | 1 (2.0) | 0 |  |
| MTX dose [mg/week], mean (SD) | 13.2 (4.3) | 13.1 (4.2) | 0.935 |
| MTX oral application, n (%) | 23 (45.1) | 3 (23.1) | 0.187 |
| Additional prednisolone, n (%) | 13 (25.5) | 7 (53.8) | 0.054 |
| Prednisolone dose [mg/day], mean (SD) | 2.8 (1.3) | 2.9 (2.0) | 0.890 |
| Vaccination |  |  | 0.336 |
| BNT162b2, n (%) | 35 (68.6) | 12 (92.3) |  |
| mRNA-1273, n (%) | 7 (13.7) | 1 (7.7) |  |
| AZD1222, n (%) | 7 (13.7) | 0 |  |
| AZD1222 + BNT162b2, n (%) | 2 (3.9) | 0 |  |
| vaccine interval in days, mean (SD) | 43 (15) | 30 (9) | **<0.001** |
| days from second vaccination, mean (SD) | 30 (22) | 39 (21) | 0.174 |
| MTX-hold, n (%) | 30 (58.8) | 1 (7.7) | **0.002** |
| for both vaccinations, n | 23 | 1 | / |
| for only the 1st vaccination, n | 2 | 0 | / |
| for only the 2nd vaccination, n | 5 | 0 | / |

\* Defined by anti-SARS-CoV-2-RBD-IgG > 1.00 S/CO.
† P values were calculated using the exact unconditional z-pooled test for binary variables (female, MTX oral application, additional prednisolone, MTX hold), Chi<sup>2</sup> test for categorical variables (medication, vaccination) and unpaired t test with Welch's correction for continuous variables.
‡ Adalimumab, Certolizumab, Etanercept, Golimumab, Infliximab.
§ Additional lowdose prednisolone for n=1.
\*\* Secukinumab.
†† Ustekinumab.
MTX, methotrexate; HCQ, hydroxychloroquine.

## Figures

**Supplementary figure 1:**
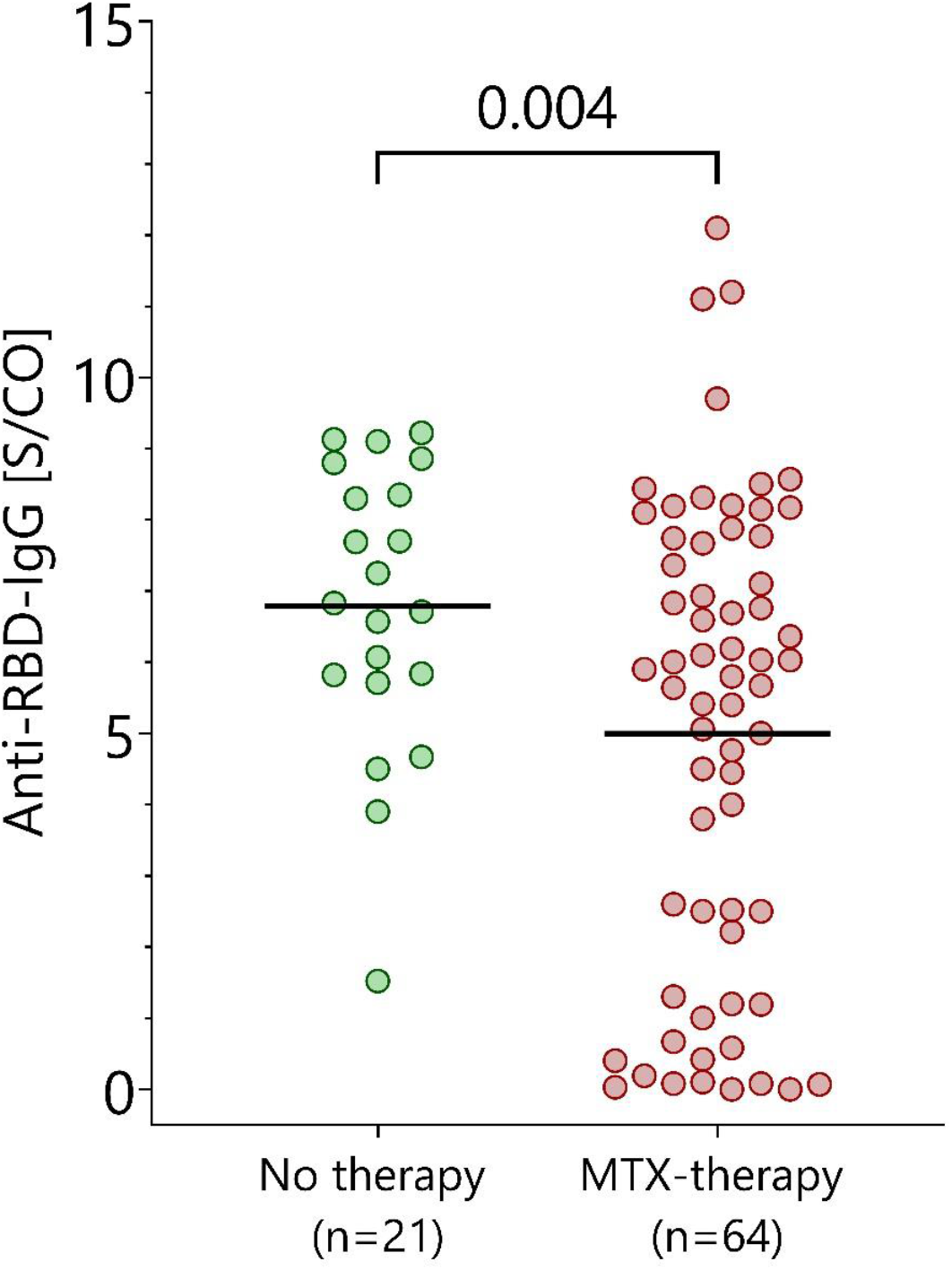
Comparison of anti-RBD-IgG-antibody levels in AIRD patients without immunosuppression and with MTX therapy. Anti-RBD-IgG-antibody levels after second vaccination in MTX-patients (n=64) represented by red dots vs. patients who were under no immunosuppressive therapy during both vaccinations (n=21) represented by green dots. P values were calculated using the parametric unpaired t test with Welch’s correction.

**Supplementary figure 2:**
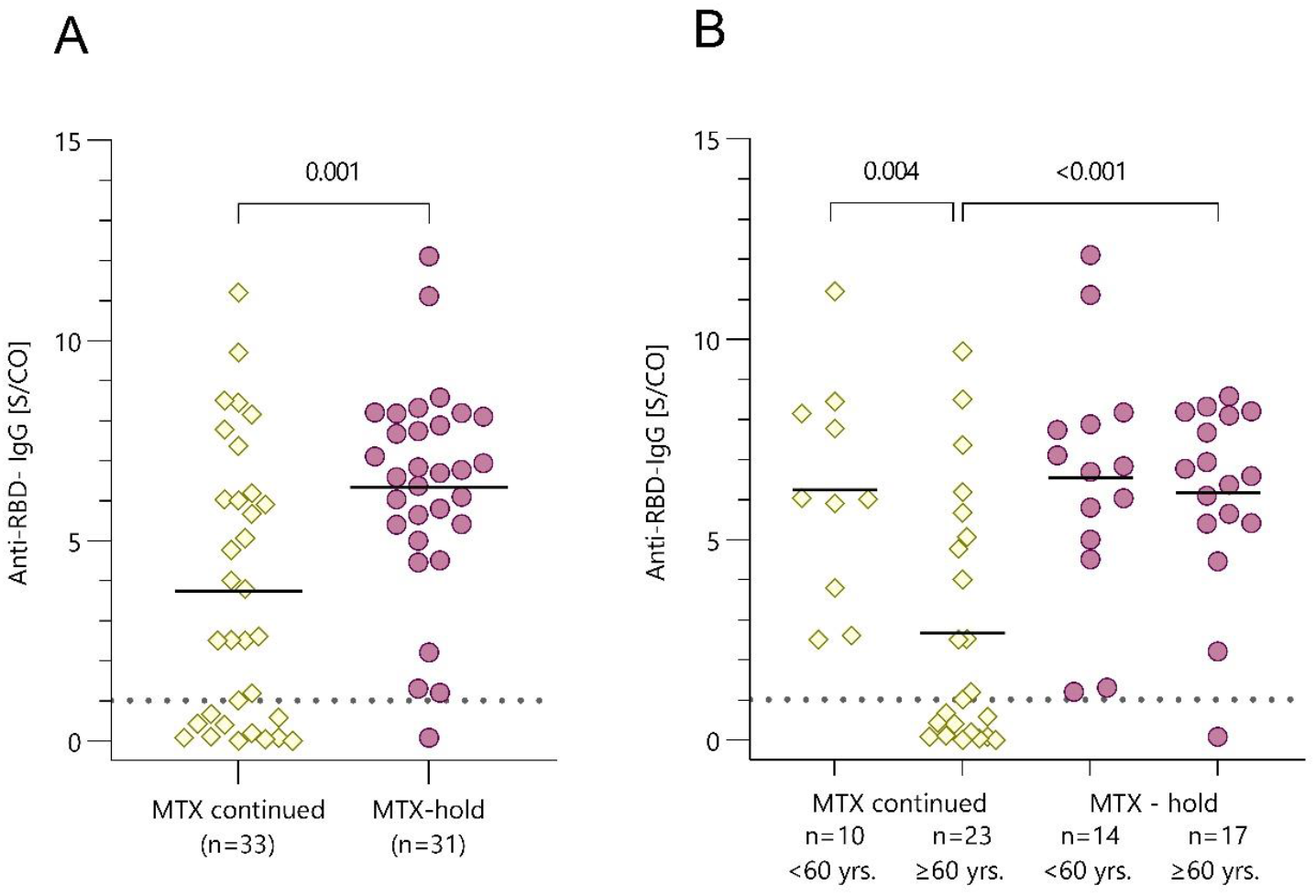
Comparison of AIRD patients who continued or held their MTX during the COVID-19 vaccination. (A) Anti-RBD-IgG concentrations compared between patients who held MTX during vaccination (n=33) and patients who continued MTX-therapy (n=31). (B) Anti-RBD-IgG concentrations differentiated by age groups <60 years and ≥60 years. P values were calculated using the parametric unpaired t test with Welch’s correction. Dotted line marks the cut-off value following manufacturer’s protocol (>1 S/CO). Yellow squares represent patients who continued MTX-therapy, purple dots represent patients who held MTX for at least one vaccination.

**Supplementary figure 3:**
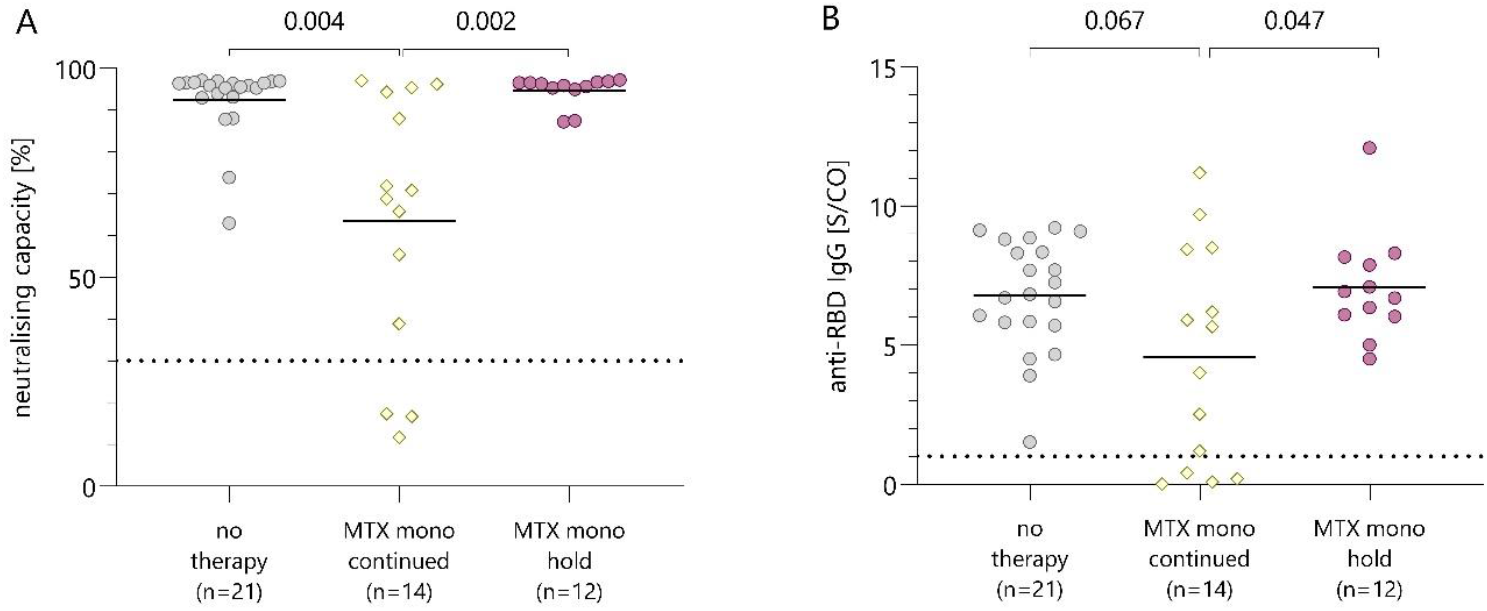
Comparison of antibody response in AIRD patients without immunosuppressive medication and MTX monotherapy patients who continued (n=14) or held (n=12) their MTX during COVID-19 vaccination. (A) Neutralising capacity measured using surrogate virus neutralisation test. Dotted line marks the cut-off value following manufacturer’s protocol (≥30 %). (B) Anti-RBD-IgG-antibody levels after second COVID-19 vaccination. Dotted line marks the cut-off antibody concentration for adequate humoral immune response following manufacturer’s protocol (>1 S/CO). P values were calculated using the parametric unpaired t test with Welch’s correction. Yellow squares represent patients who continued MTX-therapy, purple dots represent patients who held MTX for at least one vaccination.

**Supplementary figure 4:**
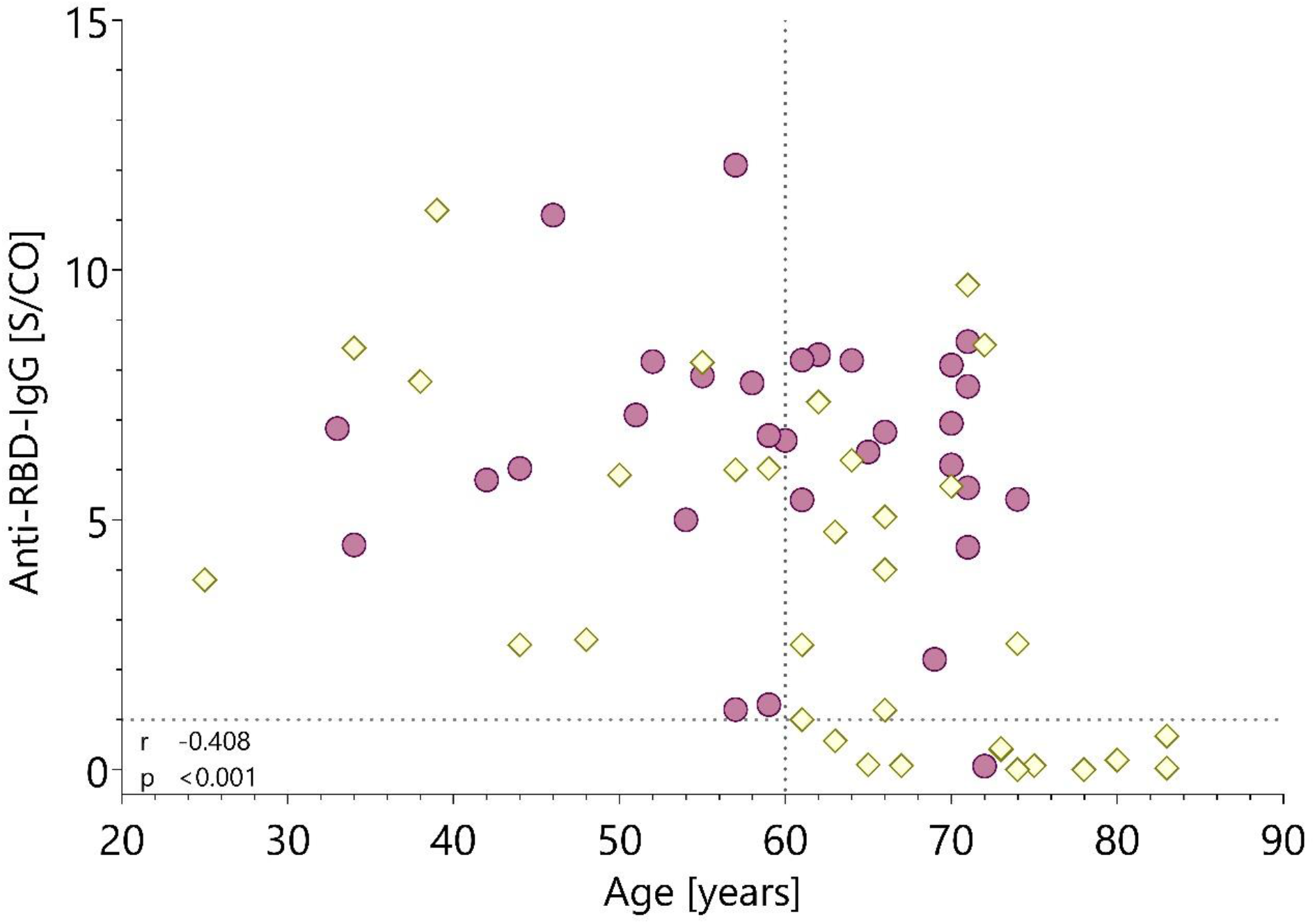
Correlation of Age and Anti-RBD-IgG-Antibody concentrations. Purple dots represent patients who held MTX during vaccination (n=31), yellow squares represent patients who continued MTX-therapy (n=33). Dotted lines mark the cut-off antibody concentration for adequate humoral immune response following manufacturer’s protocol (>1 S/CO) and the cut-off age observed for this cohort at 60 years. P value and correlation coefficient were calculated using the Spearman Rank correlation.

